# High-dimensional CyTOF profiling reveals distinct maternal and fetal immune landscapes in gestational diabetes mellitus

**DOI:** 10.64898/2026.02.17.26346459

**Authors:** Duan Ni, Felix Marsh-Wakefield, Helen M. McGuire, Angela Sheu, Xiyian Chan, Wendy Hawke, Stephanie Kullmann, Julia Sbierski-Kind, Frederic Sierro, Sue Mei Lau, Ralph Nanan

**Affiliations:** Sydney Medical School Nepean, The University of Sydney, Sydney, NSW, Australia; Charles Perkins Centre, The University of Sydney, Sydney, NSW, Australia; Nepean Hospital, Nepean Blue Mountains Local Health District, Sydney, NSW, Australia; Liver Injury & Cancer Program, Centenary Institute, Sydney, NSW, Australia; Human Immunology Laboratory, School of Medical Sciences, Faculty of Medicine and Health, The University of Sydney, Sydney, NSW, Australia; Translational Immunology Laboratory, School of Medical Sciences, Faculty of Medicine and Health, The University of Sydney, New South Wales, Australia; Department of Diabetes and Endocrinology, Prince of Wales Hospital, Sydney, NSW, Australia; School of Clinical Medicine, University of New South Wales, Sydney, NSW, Australia; Department of Obstetrics and Gynecology, Royal Hospital for Women, Randwick, Sydney, NSW, Australia; Institute for Diabetes Research and Metabolic Diseases (IDM) of Helmholtz Munich at the University of Tübingen, Tübingen, Germany; Internal Medicine IV, Department of Diabetology, Endocrinology and Nephrology, University of Tübingen, Tübingen, Germany; German Center for Diabetes Research (DZD), Tübingen, Germany; The M3 Research Center, University Clinic Tübingen (UKT), Medical Faculty, Tübingen, Germany; Health Research and Technology Group, Australian Nuclear Science and Technology Organisation (ANSTO), Sydney, NSW, Australia

**Keywords:** Gestational diabetes mellitus (GDM), cytometry by time of flight (CyTOF), maternal, fetal, cord blood, T cell, B cell

## Abstract

**Aims:** Gestational diabetes mellitus (GDM) is the most common pregnancy-related medical complication. GDM is linked to aberrant immune responses in both mothers and offsprings, specifically, the subsequent development of inflammatory diseases. Whereas prior research has focused on specific immune cell subsets, a comprehensive overview of the impacts of GDM on maternal and fetal immune landscape is lacking. Here, we aim to comprehensively decipher how GDM modulates various immune cell populations in mothers and offsprings.

**Methods:** A prospective, longitudinal case-control study was carried out. Maternal blood from GDM-affected (GDM, n=18) and non-GDM-affected (Ctrl, n=21) mothers were collected at ante-(36-38 weeks of gestation) and post-partum (6-8 weeks post-partum) timepoints. Cord blood from GDM (n=7) and Ctrl (n=11) pregnancies were collected upon C-section. They were analyzed with the state-of-the-art cytometry by time of flight (CyTOF) with a 40-marker panel. Additionally, a publicly available RNA-seq dataset for cord blood mononuclear cells was re-analyzed to confirm results from CyTOF experiments.

**Results:** Compared to Ctrl, GDM was associated with more activated maternal T cell subsets ante-partum, including increased CD45RO^+^ and Ki67^+^ CD4^+^ T cell populations, which reverted post-partum. GDM-affected maternal innate lymphoid cell (ILC) also exhibited increased granzyme B production ante-partum. On the other hand, in GDM-impacted cord blood, fetal T and B cells were more activated, displaying less naïve and more effector phenotypes, further supported by RNA-seq analyses.

**Conclusions:** Our comprehensive analyses revealed that GDM is linked to profound changes in the immune landscapes of the mothers (ante-/post-partum) and foetuses (at birth), casting novel insights towards GDM pathophysiology. Longitudinal immune profiling might be warranted for early detection and stratification of risk, and development of targeted interventions to prevent inflammatory disorders in GDM mothers and their offspring.

**Research in context:**

- What is already known about this subject?
  - *The maternal and intrauterine environments are important contributors to long-term health outcomes of mothers and offsprings*.
  - *Some maternal and fetal immunity changes have been observed in gestational diabetes mellitus (GDM)-affected pregnancies*.
  - *GDM is associated with increased risk of later-life metabolic and inflammatory diseases in mothers as well as offsprings*.
- What is the key question?
  - *What are the longitudinal alterations in maternal and fetal immune landscapes in GDM-affected pregnancies?*
- What are the new findings?
  - *High-dimensional immune profiling provided the most comprehensive overview of alterations in maternal and fetal immune landscapes associated with GDM*.
  - *GDM is associated with skewing of maternal CD4*^*+*^ *T cell and ILC towards activated phenotypes ante-partum*.
  - *GDM is linked to more activated fetal T and B cell profiles*.
- How might this impact on clinical practice in the foreseeable future?
  - *Understanding the complex alterations in the maternal and fetal immune landscape in GDM-affected pregnancy provides insights into the long-term impacts of GDM on the mother and offspring*.

## Introduction

Gestational diabetes mellitus (GDM) is among the most common complications in pregnancy, affecting more than 15% of live births worldwide [1]. GDM not only complicates pregnancy but also correlates with a plethora of long-term sequelae in affected mothers and children later in life, such as metabolic diseases like diabetes [1] and inflammatory diseases, including allergies [2] and cardiovascular diseases [3]. In this context, GDM appears to substantially modulate maternal and fetal immunity. Hence, understanding how the overall maternal and fetal immune landscapes are affected by GDM is critical for identifying and stratifying women and children at risk of GDM sequalae. It will provide insights into the mechanisms underlying these conditions, thereby informing their prevention and/or intervention. Nevertheless, previous research mainly focused on changes of specific components of the immune system. By and large, they lack comprehensive overview of the maternal and fetal immune landscapes.

We conducted a prospective, longitudinal case-control study to comprehensively analyze the immune landscapes of the mothers (ante-/post-partum) and foetuses (at birth), comparing GDM-affected (GDM) with non-GDM-affected pregnancies (Ctrl). Harnessing the state-of-the-art high-dimensional cytometry by time of flight (CyTOF) technique, we found that GDM disrupted T cell, B cell and innate lymphoid cell (ILC) compartments. Specifically, GDM was associated with a more activated maternal T cell and ILC phenotype ante-partum, which reverted post-partum. Additionally, GDM was linked to fetal T cell and B cell activation. Together, our unprecedented analyses provide in-depth insights towards changes in maternal and fetal immune landscapes associated with GDM.

## Methods

### Cohort

This prospective longitudinal study was based in the Royal Hospital for Women, Sydney. GDM was diagnosed using the IADPSG 2010 criteria (fasting glucose ≥5.1 mmol/L, 1 hour glucose ≥8.5 mmol/L and/or 2-hour glucose ≥10.0 mmol/L on universal glucose tolerance screening test). Mothers with GDM (n=18) were recruited in their third trimester from the GDM clinic. Non-GDM Ctrl mothers (n=21) were recruited in their third trimester in the antenatal clinic. Exclusion criteria included hypertensive disorders and a history of inflammatory or autoimmune diseases [4]. This study was approved by the South Eastern Sydney Local Health District Human Research Ethics Committee (12/271) and all study participants provided informed consent.

Clinical characteristics and timeline of the study are described in Figure S1. Maternal blood was collected at 36-38 weeks of gestation and not during labour or delivery (ante-partum, hereafter refer to as *37w* timepoint) and 6-8 weeks post-partum (*ppm* timepoint); and venous cord blood was collected at the time of elective C-section.

### Cytometry by time of flight (CyTOF)

Blood samples were processed as previously described [4] to obtain peripheral blood mononuclear cells (PBMCs, maternal) and cord blood mononuclear cells (CBMCs, fetal).

Cryopreserved PMBCs and CBMCs were thawed in 10% FBS supplemented RPMI-1640 (Gibco, UK) containing 1:10,000 universal nuclease (Thermo Fisher Waltham, MA) and stained with 1.25 uM Cell-ID cisplatin (Standard BioTools) in PBS for 3 min at room temperature. Cells were then stained with a cocktail of surface marker antibodies (Table S1) in CyTOF staining buffer (PBS with 0.5% bovine serum albumin [BSA; Sigma] and 0.02% sodium azide [Sigma]) for 30 min at 4°C. After surface staining, cells were washed with CyTOF staining buffer and then fixed and permeabilized using eBioscience’s FoxP3 buffer kit (San Diego, CA, USA) at 4°C for 45 min and stained with intracellular antibodies for 30 min on ice. Cells were then washed twice and fixed in 4% paraformaldehyde containing DNA intercalator (0.125 uM Iridium-191/193; Standard BioTools). Prior to data acquisition, cells were washed by centrifugation in CyTOF staining buffer, then ultrapure water, filtered through a 35_μ_m nylon mesh and diluted to 800,000 cells/ml in MilliQ with 1/10 dilution of 4-Element EQ normalization beads (Standard BioTools). Cells were acquired at a rate of 200-400 cells/s using a CyTOF 2 Helios upgraded mass cytometer (Standard BioTools, Toronto, Canada).

Samples were stained and run in 6 batches. Matched samples from the same individual were run in the same batch. Each batch also contained a batch control consisting of one replicate aliquot of PBMC from a healthy donor, thawed and stained in parallel with study samples.

Cells were normalized for signal intensity of EQ beads using the Helios software. FlowJo v10.9.0 software (BD Biosciences, NJ, USA) was used to gate populations of interest based on the gating strategy in Figure S2-5. Samples with fewer than 3000 cells were excluded from analysis due to low sample quality.

### Statistical and bioinformatic analyses

Maternal results were compared with mixed effect analyses with time (37w vs ppm) and GDM status (Ctrl vs GDM) as factors based on their pairing at two timepoints. Fetal results were analyzed with an unpaired t-test. P<0.05 was considered statistically significant. These analyses were run in GraphPad PRISM.

For Gene Set Enrichment Analysis (GSEA), a CBMC RNA-seq normalized dataset (GSE203346) was downloaded from the original publication [5] and analyzed with GSEA following the developer’s tutorial based on the human Hallmark Gene Sets, which cover 50 representative gene sets involved in most cellular and molecular activities.

## Results

### Study overview

As in Figure S1A, blood samples were collected from pregnant women around 36-38 weeks of gestation (ante-partum, referred to as *37w* hereafter, 21 healthy control (Ctrl) and 18 GDM), and 6-8 weeks post-partum (referred to as post-partum, *ppm*, 9 Ctrl and 12 GDM), respectively. 6 Ctrl and 9 GDM mothers had samples collected at both timepoints. Cord blood samples were available from 11 Ctrl and 7 GDM-impacted pregnancies upon C-section. Clinical characteristics were comparable in GDM and Ctrl groups, except glucose tolerance test results (Figure S1B-C).

PBMCs/CBMCs were subject to high-dimensional CyTOF analysis with a 40-color panel (Table S1), which comprehensively profiled major immune cell populations (T, B, natural killer (NK), NKT cells, innate lymphoid cells (ILCs), monocytes, conventional dendritic cells (cDCs), and plasmacytoid dendritic cells (pDCs)) following the gating strategy in Figure S2-5.

### Maternal immune profiles

Overall, major immune cell populations in maternal blood were mildly affected by GDM ante- and post-partum (Figure 1A), with the exception of total ILCs, which were higher in Ctrl compared to GDM mothers ante-partum (37w). Our longitudinal sampling allowed for direct comparisons before and after delivery (ante-vs post-partum). At post-partum timepoint, whilst T cells expanded, classical and intermediate monocytes decreased in GDM-affected mothers. In contrast, total ILCs decreased in Ctrl, to an equivalent proportion as post-partum GDM (Figure 1A).

**Figure 1.**
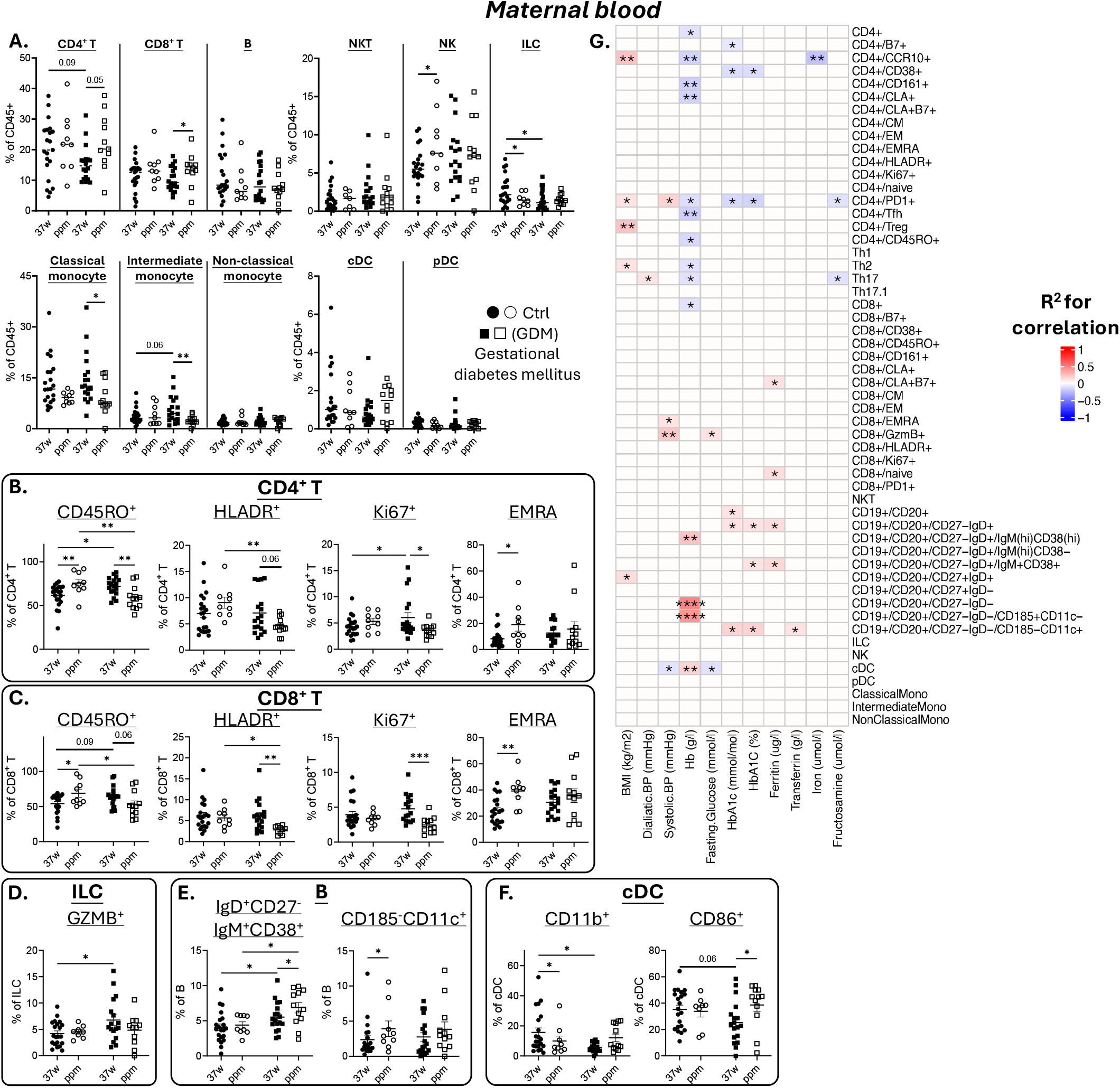
An overview of the changes in the maternal immune landscape from mothers with (GDM, marks in square) or without (Ctrl, marks in circle) GDM at 36-38 weeks of gestation (ante-partum, 37w, filled marks) and post-partum (ppm, hollow marks) timepoints. **A**.Changes of major immune cell populations (CD4^+^ T cell, CD8^+^ T cell, B cell, natural killer T (NKT) cell, natural killer (NK) cell, total innate lymphoid cell (ILC), classical monocyte, intermediate monocyte, non-classical monocyte, conventional dendritic cell (cDC), plasmacytoid dendritic cell (pDC)). **B-C**. Changes of major CD4^+^ T cell (**B**) and CD8^+^ T cell (**C**) subsets (CD45RO^+^, HLADR^+^, Ki67^+^, effector memory re-expressing CD45RA (EMRA)). **D**. Changes of granzyme B (GZMB)-expressing total ILC subsets (GZMB^+^). **E**. Changes of CD38^+^ na#x00EF;ve B cell (IgM^+^CD38^+^IgD^+^CD27^-^) and CD185^-^CD11c^+^ B cell subsets. **F**. Changes of activated cDC subsets (CD11b^+^, CD86^+^). **G**. Correlation analyses for the frequencies of maternal blood immune cell subsets at ante-partum timepoint with their corresponding clinical parameters. The pseudo-colour gradient reflects the R^2^ for correlation analysis. Grids in red and blue denote positive and negative correlations respectively, and grids in white mean no significant correlation. (data are represented as mean ± S.E.M., with ^*^*p* < 0.05, ^**^*p* < 0.01, ^***^*p* < 0.001, ^****^*p* < 0.0001)

We next examined in detail the subsets of different maternal immune cell populations. During pregnancy, GDM was associated with maternal CD4^+^ T cells skewed towards activation, as suggested by their increased CD45RO^+^ (effector function) and Ki67^+^ (proliferation) population frequencies (Figure 1B). Interestingly, these changes reverted post-partum, as in healthy controls, CD4^+^ and CD8^+^ T cells exhibited higher proportions of CD45RO^+^ and HLADR^+^ (activation) subsets (Figure 1B-C). Longitudinally, in GDM group, percentages of T cells expressing activation markers CD45RO, HLADR, and Ki67 consistently decreased post-partum. Contrarily, in Ctrl, EMRA (effector memory re-expressing CD45RA) and CD45RO^+^ T cells expanded after delivery (Figure 1B-C). Similar to T cells, ILCs from GDM mothers exhibited an activated phenotype ante-partum, as more of them expressed granzyme B (Figure 1D). B cell subsets were minimally impacted by GDM, except for proportions of CD38^+^ naïve B cells (IgM^+^CD38^+^IgD^+^CD27^-^) among total B cells, which were consistently higher in GDM mothers both ante- and post-partum. Frequencies of IgD^-^CD27^-^CD185^-^CD11c^+^ B cells were increased after delivery in Ctrl but not in GDM-affected mothers (Figure 1E). cDCs appeared to be more activated in Ctrl at 37w, with higher proportions of cells expressing activation markers CD11b and CD86 (Figure 1F), which did not persist postpartum. No difference was found for other myeloid cell subsets.

To further interrogate the relationships between metabolic health and maternal immune profiles ante-partum, we performed correlation analyses for both Ctrl and GDM mothers, comparing frequencies of major immune cell subsets with various clinical and metabolic parameters (Figure 1G). Maternal body mass indexes (BMI) positively correlated with several T cell subsets. In contrast, hemoglobin (Hb) and HbA1c levels were linked to decreased T cell population frequencies but increased B cell subset proportions. Fasting glucose levels were linked to higher granzyme B-expressing CD8^+^ cytotoxic T cell frequencies but lower cDCs (Figure 1G).

### Fetal immune profiles

To probe the impacts from GDM on fetal immune profiles, we analyzed CBMCs using a similar approach, covering more than 50 major immune subsets. In GDM-affected cord blood, there were higher frequencies of CD4^+^ and CD8^+^ EM (effector memory), CD4^+^ EMRA, CD8^+^ CM (central memory), CD8^+^CLA^+^ (skin-homing CD8^+^ T cells) and CD4^+^Ki67^+^ cells among total lymphocytes. In Ctrl group, frequencies of total monocytes, classic monocytes, total B cells, naïve B cells, and IgM^+^CD38^+^ naïve B cells were higher (Figure 2A). Cord blood T cells from GDM group also appeared to be more activated, reflected by their less naïve but more effector memory-like phenotypes. There were also marginally more CD8^+^ T cells producing granzyme B in GDM group (Figure 2B-C). Among B cells, GDM group had higher proportions of CD38^-^ naïve B cells (IgM^hi^CD38^-^IgD^+^CD27^-^) and IgD^+^CD27^+^ unswitched memory B cells (activated B cells that have not gone through IgM class switch) (Figure 2D). Other populations in CBMCs including NK cells, DCs and CD34^+^ hematopoietic stem cells (HSCs) showed no difference between Ctrl and GDM groups (Figure 2A).

**Figure 2.**
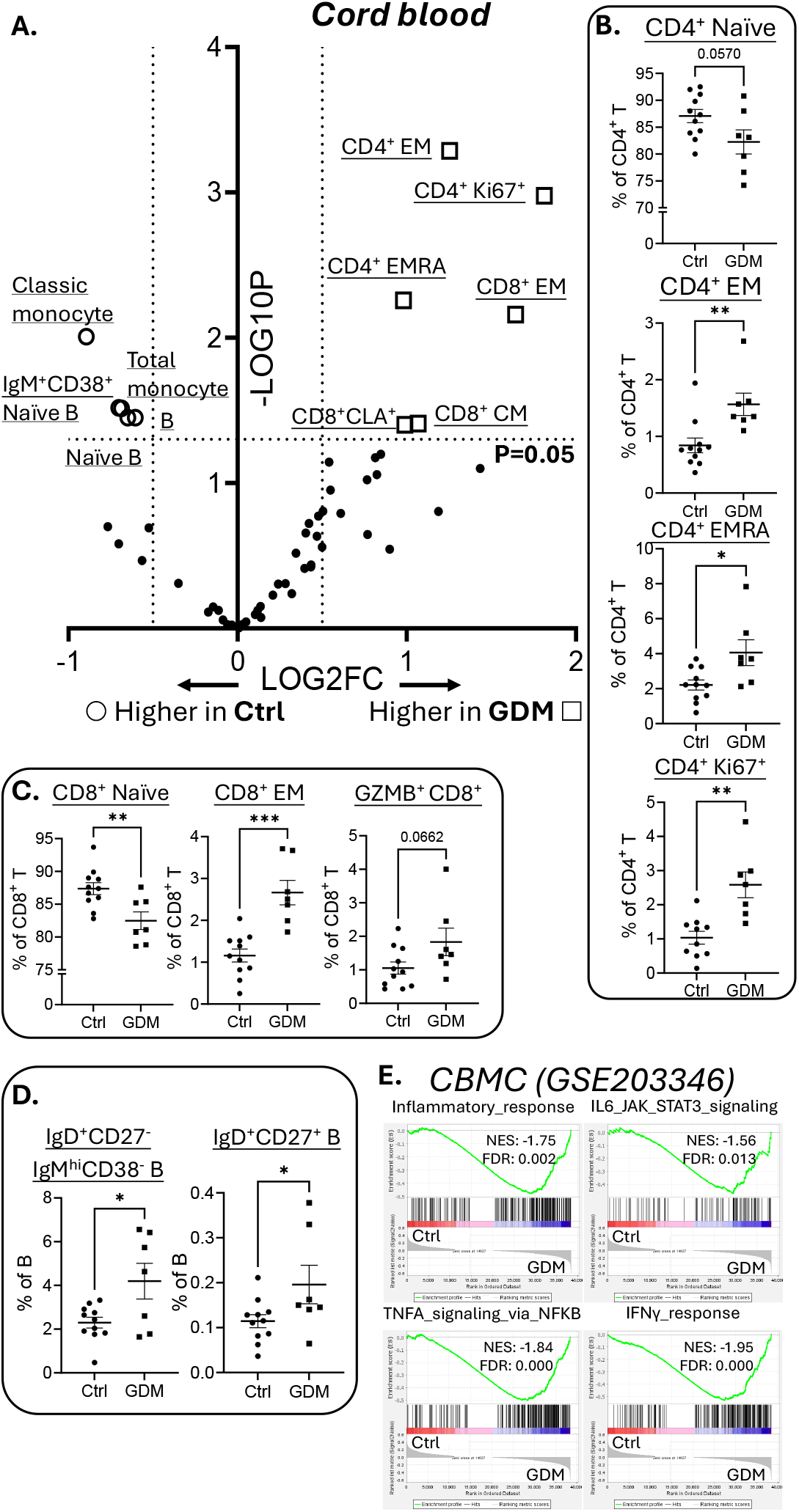
An overview of the changes in the fetal immune landscape from pregnancies affected (GDM, marks in square) versus not affected (Ctrl, marks in circle) by GDM. A.Volcano plot comparing the changes in frequencies of different fetal immune cell subsets. **B**. Changes of major CD4^+^ T cell subsets (naïve, effector memory (EM), EMRA, Ki67^+^). **C**. Changes of major CD8^+^ T cell subsets (naïve, EM, GZMB^+^) **D**. Changes of major B cell subsets (CD38^-^ naïve B cells (IgM^hi^CD38^-^IgD^+^CD27^-^), IgD^+^CD27^+^ unswitched memory B cell) **E**. Gene set enrichment analysis results comparing the transcriptomic profiles of cord blood mononuclear cells (CBMCs) in “Inflammatory responses”, “IL6-JAK-STAT3 signaling”, “TNF_α_ signaling via NF_κ_B” and “IFN_γ_ response” gene sets based on dataset GSE203346. (data are represented as mean ± S.E.M., with ^*^*p* < 0.05, ^**^*p* < 0.01, ^***^*p* < 0.001, NES: normalized enrichment score, FDR: false discovery rate)

To further corroborate our findings, an RNA-seq dataset (GSE203346) for CBMCs from Ctrl or GDM-affected pregnancies was re-analyzed with Gene Set Enrichment Analysis (GSEA). GSEA found that GDM-impacted CBMCs were enriched in several inflammation-related gene sets (Figure 2E), aligned with the more activated/effector-like phenotypes of CBMC T cells unveiled by CyTOF in our study.

## Discussion

Here, we presented comprehensive analyses of the maternal and fetal immune landscapes in GDM in a longitudinal cohort. Utilizing the state-of-the-art CyTOF platform, our high-dimensional analysis covered 40 markers and captured more than 50 major immune cell subsets, surpassing most prior studies that generally only focused on specific immune compartments [6]. Moreover, longitudinal analyses on ante- and post-partum maternal samples offered additional insights into the long-term influences from GDM, compared to previous reports that typically examined a single timepoint during pregnancy or GDM manifestation. Importantly, we also unveiled detailed changes in cord blood immune profiles, especially in the T and B cell compartments.

We found that GDM compared to non-GDM Ctrl during pregnancy was linked to maternal immune activation, particularly within T cell compartments, while the inverse was observed post-partum. Given that parturition is typically accompanied with maternal metabolic shift, including resolution of gestational insulin resistance [7], it remains to be determined whether such metabolic recovery might mediate the observed post-partum remodelling of the maternal immune landscapes, which might be linked to the development of non-communicable diseases, as frequently reported in women with GDM [1].

In contrast to T cells, GDM had limited effects on other maternal PBMC populations. Notably, GDM was associated with lower total ILC frequency. Given the critical roles of ILCs in maintaining metabolic and tissue homeostasis [6, 8], more in-depth explorations of different ILC subsets and their functionality would be of interests.

Aligning with our previous findings [9], we observed immune activation, particularly in T cells, in cord blood from GDM-affected pregnancies. Such immune imprinting may contribute to the previously reported associations between GDM and an increased risk of T cell-mediated inflammatory conditions in offspring, such as allergic diseases [1, 2]. However, the mechanisms underlying this elevated activation state and its persistence in longer term remain to be elucidated.

On the other hand, GDM’s influences on cord blood B cell populations are generally lacking previously. This is addressed in our present study, where we revealed some notable alterations in cord blood B cell compartments associated with GDM. In addition to lower total B cell frequency, GDM seemed to correlate with B cell activation, as reflected by lower naïve but higher activated phenotypes (IgD^+^CD27^+^). This aligns with our previous reports using scRNA-seq that GDM cord blood B cell had a more inflamed transcriptomic profile [9], similar to aforementioned findings in T cell compartments. These results suggest GDM might rewire the offsprings’ immune system towards an inflammatory status, potentially contributing to their increased susceptibility to non-communicable diseases [1, 2].

A prior study documented that GDM might dampen CB HSC differentiation towards the lymphoid lineage but promote development into erythroid progenitors [10]. Whether this might explain our above observations regarding T and B cells warrant further investigations, though our CyTOF analysis found comparable HSC frequencies in Ctrl and GDM groups.

Collectively, our comprehensive immune landscape analyses revealed the multifaceted GDM-associated changes in both maternal and fetal immunity. In the context of crosstalk between metabolism and immunity, these findings might provide new insights into the pathophysiology of GDM-related pregnancy complications and their long-term sequelae. With further validations in larger and duration-extended cohorts, our approaches and findings could inform future strategies for monitoring and managing GDM and its sequelae in mothers and their offsprings.

## Data Availability

All data produced in the present study are available upon reasonable request to the authors

## Abbreviations

BMI: body mass index
CBMC: cord blood mononuclear cell
cDC: conventional dendritic cell
CyTOF: cytometry by time of flight
EM: effector memory
EMRA: effector memory re-expressing
CD45RA FDR: false discovery rate
Granzyme B: GZMB
GDM: gestational diabetes mellitus
GSEA: Gene Set Enrichment Analysis
Hb: hemoglobin
HSC: hematopoietic stem cell
ILC: innate lymphoid cell
NES: normalized enrichment score
NK cell: natural killer cell
PBMC: peripheral blood mononuclear cell
pDC: plasmacytoid dendritic cell
Ppm: post-partum

## Acknowledgements

The authors thank Dr. Ellis Patrick and Dr. Caryn van Vreden for their helpful discussions in CyTOF experiments. The authors would like to thank all the participants involved in this study.

## Ethics statement

This study was approved by the South Eastern Sydney Local Health District Human Research Ethics Committee (12/271) and all study participants provided informed consent.

## Contribution statement

Conception of the study: D.N., F.S., S.M.L., & R.N.; clinical study: F.S. & S.M.L.; experiment, data acquisition and analyses: D.N., F.M-W., H.M.M., & R.N.; manuscript drafting: D.N., F.S., & R.N.; critical revision of the manuscript: all authors.

All authors approved the final version of the manuscript.

