## Supplementary Information for "High-dimensional CyTOF profiling reveals distinct maternal and fetal immune landscapes in gestational diabetes mellitus"

### Figure S1.

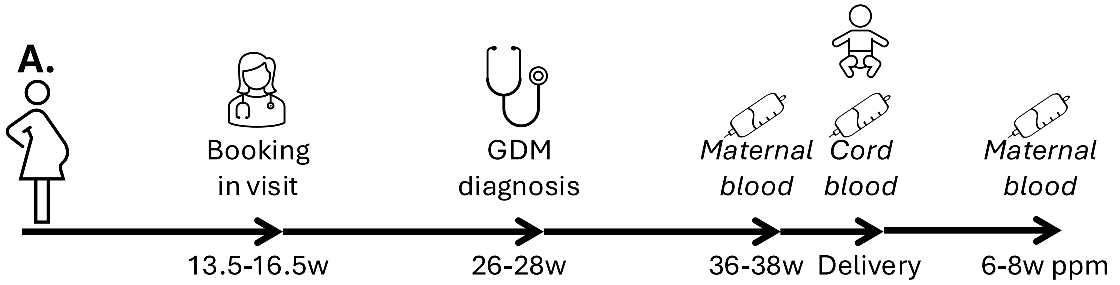


**B.**

| *Maternal blood samples* | 37w timepoint | | | Post-partum timepoint | | |
|---|---|---|---|---|---|---|
| | ***Ctrl*** | ***GDM*** | **p value** | ***Ctrl*** | ***GDM*** | **p value** |
| **n** | 21 | 18 | - | 9 | 12 | - |
| **Age, years** | 33.24±1.205 | 34.61±0.8407 | 0.3715 | 32.89±1.968 | 34.33±1.170 | 0.5134 |
| **Ancestral background** | 16 Caucasian, 3 SE Asian, 1Mediterranean, 1 other | 9 Caucasian, 2 South Asian, 5 SE Asian, 1 Mediterranean, 1 other | 0.0708 | 4 Caucasian, 2 South Asian, 1 SE Asian, 1 Mediterranean, 1 other | 7 Caucasian, 3 SE Asian, 1Mediterranean, 1 other | 0.9371 |
| **Booking BMI (kg/m2)** | 25.98±2.014 | 24.15±1.111 | 0.454 | 24.98±1.660 | 24.59±1.309 | 0.8548 |
| **Body weight (kg, booking)** | 71.52±6.033 | 65.85±3.043 | 0.4078 | 68.06±4.494 | 67.50±4.045 | 0.9284 |
| **Body weight (kg, GDM diagnosis/Ctrl 24-28w)** | 84.61±7.826 | 72.39±3.112 | 0.1083 | 81.98±5.419 | 73.47±4.291 | 0.2676 |
| **Booking systolic BP (mmHg)** | 105.4±2.182 | 110.4±2.510 | 0.1371 | 109.2±2.012 | 111.6±3.163 | 0.5447 |
| **Booking diastolic BP (mmHg)** | 64.9±1.416 | 66.69±1.683 | 0.4207 | 67.78±1.949 | 69.10±2.584 | 0.6932 |
| **Delivery by c-section, n (%)** | 17 (80.95%) | 10 (55.56%) | 0.0867 | 4 (44.44%) | 5 (41.66) | 0.1273 |
| **Diagnostic GTT** | | | | | | |
| **Fasting glucose (mmol/l)** | 4.295±0.06176 | 4.835±0.1483 | **0.0011** | 4.367±0.06667 | 4.736±0.1712 | 0.0803 |
| **1h glucose (mmol/l)** | 6.833±0.2772 | 9.117±0.6102 | **0.0007** | 6.800±0.2843 | 9.817±0.3146 | **<0.0001** |
| **2h glucose (mmol/l)** | 5.405±0.2199 | 8.476±0.4290 | **<0.0001** | 5.811±0.2214 | 8.736±0.3971 | **<0.0001** |
| **Insulin required?** | - | 16Y 2N | - | - | 12Y | - |
| **Metformin started?** | - | 1Y 17N | - | - | 1Y 11N | - |
| **HbA1c (%)** | 5.210±0.09708 | 5.367±0.09532 | 0.259 | 5.211±0.1829 | 5.275±0.1045 | 0.7511 |
| **HbA1c (mmol/mol)** | 33.48±1.048 | 35.11±1.013 | 0.2733 | 33.67±1.929 | 34.17±1.107 | 0.8141 |
| **Fructosamine (μmol/l)** | 187±3.309 | 193.6±3.744 | 0.1948 | 188.6±4.652 | 191.2±2.949 | 0.6255 |
| **Iron (umol/l)** | 19.26±1.213 | 18.77±2.123 | 0.8345 | 17.50±2.341 | 17.93±2.516 | 0.9042 |
| **Transferrin (g/l)** | 3.962±0.1092 | 4.153±0.1873 | 0.3632 | 3.733±0.1878 | 4.291±0.2429 | 0.0969 |
| **Ferritin (μg/l)** | 36.71±6.047 | 32.65±4.317 | 0.6039 | 33.44±5.783 | 30.36±5.739 | 0.7124 |
| **Hb (g/l)** | 124.5±2.960 | 123.9±1.814 | 0.8735 | 121±2.392 | 123.8±2.720 | 0.4745 |
| **Postpartum GTT** | | | | | | |
| **Fasting glucose (mmol/l)** | - | - | - | - | 4.642±0.1177 | - |
| **1h glucose (mmol/l)** | - | - | - | - | 7.867±0.5746 | - |
| **2h glucose (mmol/l)** | - | - | - | - | 7.067±0.5447 | - |

**C.**

| *Cord blood samples* | ***Ctrl*** | ***GDM*** | **p value** |
|---|---|---|---|
| **n** | 11 | 7 | - |
| **Gravida** | 2.909±0.3921 | 3±0.3086 | 0.8714 |
| **Parity** | 1.182±0.2264 | 0.7143±0.2857 | 0.2172 |
| **Birthweight (g)** | 3594±99.19 | 3564±211.8 | 0.8896 |
| **Length (cm)** | 51.46±0.4922 | 52.57±1.232 | 0.3493 |
| **BMI** | 13.55±0.2587 | 12.88±0.5817 | 0.2497 |
| **Gender** | 8M 3F | 4M 3F | 0.6267 |

**Figure S1.** Overview of the demographic background information of the cohort and samples. (GDM: gestational diabetes mellitus; BMI: body mass index; BP: blood pressure; GTT: glucose tolerance test; Hb: hemoglobin)

Figure S2.

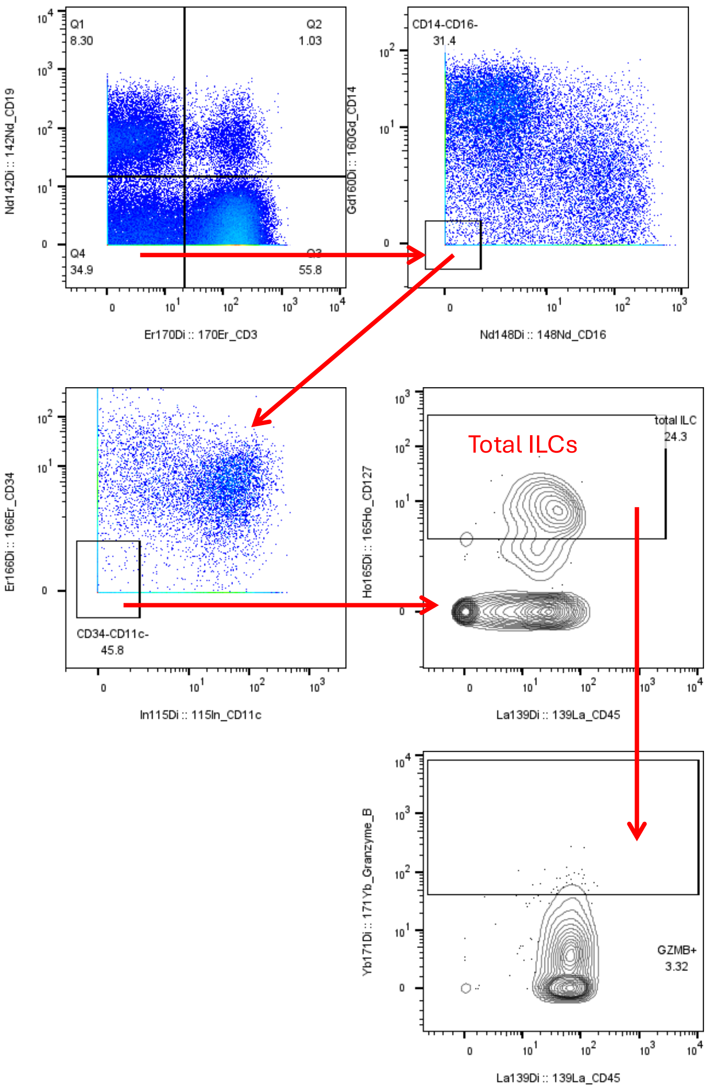


**Figure S2.** CyTOF gating strategies for innate lymphoid cells (ILCs) analysis.

Figure S3.

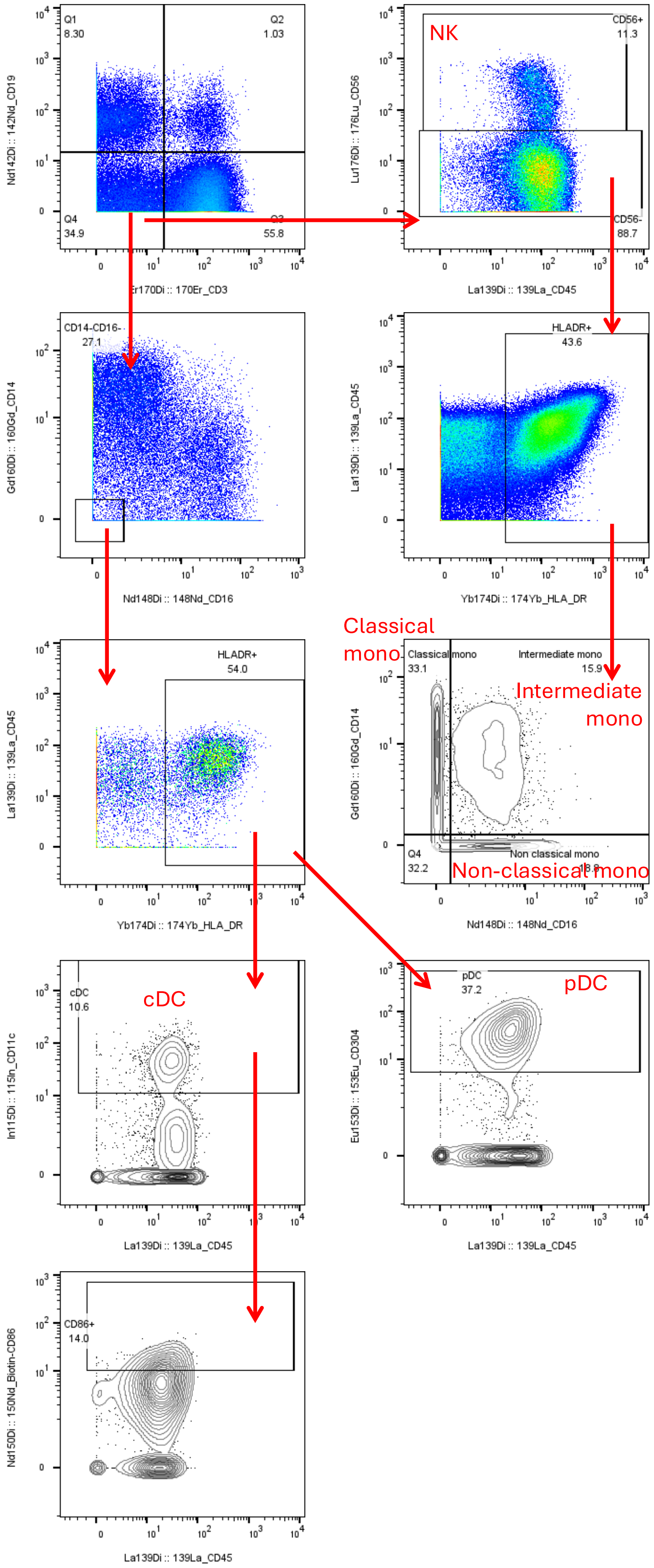


**Figure S3.** CyTOF gating strategies for myeloid cells and natural killer (NK) cells analysis.

Figure S4.

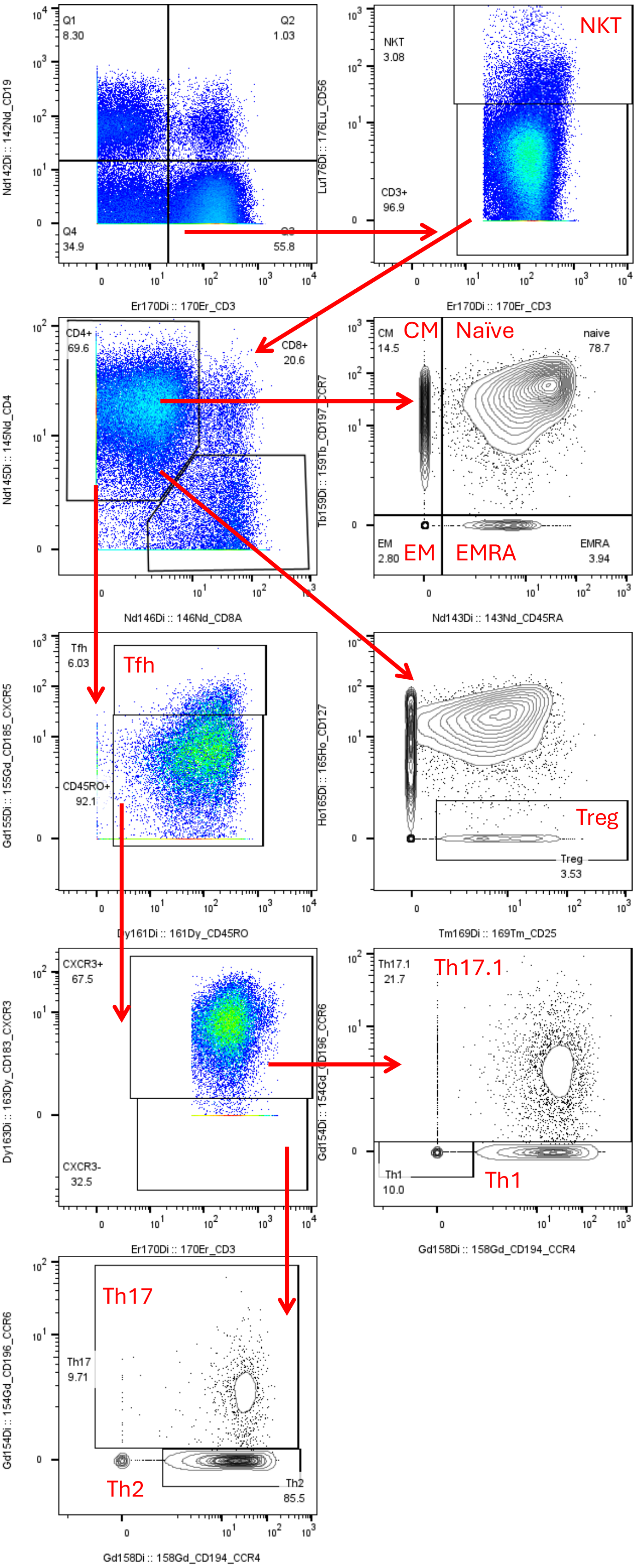


**Figure S4.** CyTOF gating strategies for T cells analysis.

Figure S5.

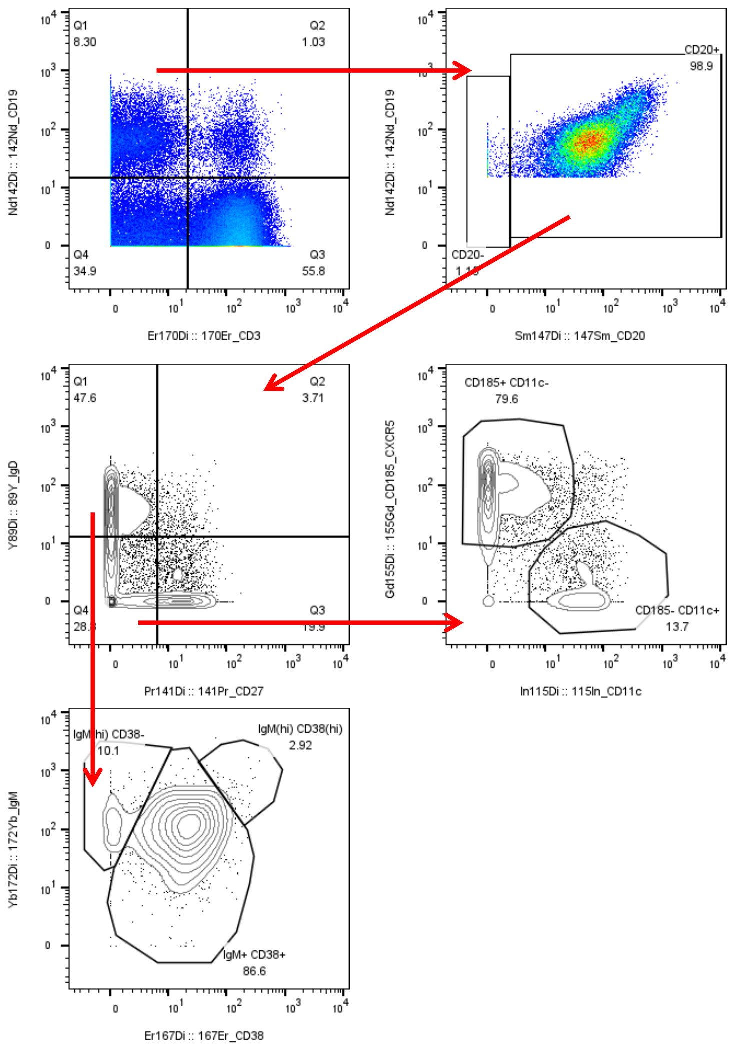


**Figure S5.** CyTOF gating strategies for B cells analysis.

**Table S1.** Antibodies used in CyTOF experiments.

| Metal label | Specificity | Antibody Clone | Manufacturer |
|---|---|---|---|
| **89Y** | IgD | IA6-2 | BD Biosciences |
| **139La** | CD45 | H130 | Biolegend |
| **115In** | CD11c | Bu15 | Biolegend |
| **141Pr** | CD27 | M-T271 | BD Biosciences |
| **142Nd** | CD19 | HIB19 | Biolegend |
| **143Nd** | CD45RA | H1100 | Biolegend |
| **144Nd** | CD195 (CCR5) | HEK/1/85a | Biolegend |
| **145Nd** | CD4 | RPA-T4 | BD Biosciences |
| **146Nd** | CD8 | RPA-T8 | Biolegend |
| **147Sm** | CD20 | 2H7 | Biolegend |
| **148Nd** | CD16 | 3GB | BD Biosciences |

| | | | |
|---|---|---|---|
| **149Sm** | CLA | HECA-452 | Biolegend |
| **Biotin** | CD86 | IT2.2 | BD Biosciences |
| **150Nd** | Biotin | 1D4-C5 | Standard BioTools |
| **151Eu** | FCRL3 | 546828 | R&D Systems |
| **152Sm** | CD117 | 104D2 | Biolegend |
| **153Eu** | CD304 | AD5-17F6 | Miltenyi |
| **154Sm** | CD196 (CCR6) | 11A9 | BD Biosciences |
| **155Gd** | CD185 (CXCR5) | RF8B2 | BD Biosciences |
| **PE** | CCR10 | 1B5 | BD Biosciences |
| **156Gd** | PE | PE001 | Biolegend |
| **158Gd** | CD194 (CCR4) | L291H4 | Biolegend |
| **159Tb** | CD197 (CCR7) | G043H7 | Biolegend |
| **160Gd** | CD14 | M5E2 | BD Biosciences |
| **161Dy** | CD45RO | UCHL1 | Biolegend |
| **162Dy** | FoxP3 | PCH101 | eBioscience |
| **163Dy** | CD183 (CXCR3) | G025H7 | Standard BioTools |
| **164Dy** | CD161 | DX12 | BD Biosciences |
| **165Ho** | CD127 | A019D5 | Biolegend |
| **166Er** | CD34 | 581/CD34 | BD Biosciences |
| **167Er** | CD38 | HIT2 | Biolegend |
| **168Er** | Ki67 | B56 | BD Biosciences |
| **169Tm** | CD25 | 2A3 | Biolegend |
| **170Er** | CD3 | UCHT1 | BD Biosciences |
| **171Yb** | Granzyme B | GB-11 | Acris |
| **172Yb** | IgM | G20 127 | BD Biosciences |
| **173Yb** | Integrin β7 | FIB504 | BD Biosciences |
| **174Yb** | HLA-DR | L243 | BD Biosciences |
| **175Lu** | CD279 (PD-1) | EH12.2H7 | Biolegend |
| **176Yb** | CD56 | NCAM16.2 | BD Biosciences |
| **209Bi** | CD11b | ICRF44 | Biolegend |
